# Non-pharmacological management of Alzheimer’s disease a qualitative study, in Mauritius

**DOI:** 10.1101/2020.01.25.20018747

**Authors:** Smita Goorah, Geeta Devi Dorkhy

## Abstract

**Introduction:** The major form of dementia is Alzheimer’s disease (AD). In Mauritius it was estimated in 2015 that around 10,000 persons had Alzheimer’s disease from Alzheimer disease International report^1^. This represent 16% of the total population and it is expected to rise. Therefore, apart from pharmacological therapies, non-pharmacological treatment (NPT) which can improve cognitive performance such as cognitive training, occupational therapy, reminiscence therapy, are being highly investigated.

**Aim and objective:** To determine the outcome(s) and to highlight the effective strategies in the management of Alzheimer’s Disease in Mauritius using NPT. Information obtained from this research will add to the current knowledge and expansion.

**Materials and methods:** The only NGO in Mauritius is “Alzheimer’s Association”, from where the persons with AD were recruited. Non-interventional methods of qualitative study involving interviews of person with AD, lasting 30-45 minutes were audio-recorded. An informed consent from the carer or patient-relative prior to starting was ensured. Both semi-structured interviews and open-ended questions were used, relating to NPT. Thematic analysis in qualitative research, followed by coding and decoding raw data. Similar categories of data were grouped and themes emerged.

**Results:** In-depth responses of the person with AD related with their experiences, perceptions, opinions, feelings, and knowledge. Qualitative study (n=20) emerged with 6 main themes. Among these, dependency on family identified as an important theme. It has also been associated as an important form of cognitive and functional engagement for person with AD.

**Discussion:** Non-pharmacological interventions has an important role to prolong the active age of older individuals, as well as to maintain quality of life.

**Conclusion:** There is still a greater demand on the market for non-pharmacological interventions and Mauritius should plan and devise national guidelines to deal with this unmet proportion of elderly so as to curb for future health equality and stability.

## INTRODUCTION

No drug can cure Alzheimer’s disease (AD) but non-pharmacological therapy (NPT) has shown to slow down the process till we have a cure^2^. (Nelson L, 2015). AD is still a chronic and serious condition especially since it is the third cause of death in year 2016, in high income countries according to World Health Organization^3^. (WHO, 2018) Therefore, any intervention which can delay the onset of symptoms in AD/ dementia will have a big impact on patients and their families.

So far, no research has been done on the Alzheimer patient in Mauritius, little is known on the NPT, therefore to be able to better understand the current situation this project was conducted with the population of Alzheimer’s patient at the NGO at Belle Rose in Mauritius.

Many side effects of drug therapy for AD is known in patients from the use of Memantine, Rivastigmine, which are acetylcholine esterase inhibitors. The non-pharmacological management was most interesting topic to find out the different outcomes and various aspects into the management of person with Alzheimer’s disease.

## MATERIALS AND METHODS

Qualitative research is a non-statistical inquiry. It is based on the analysis of a social phenomenon affecting the lives of the individual or as part of their lived-experience. Therefore, qualitative researches have an interest into developing new concepts and try to develop the power to understand theories or a hypothesis. Hence qualitative research is to understand the people from their own perspectives and their own frames of reference, how they live their daily lives, experiencing reality^4^ (Corbin & Strauss, 2008). “Grounded theory is a hypothesis-generating analytic technique whereby raw interview or observation data are systematically analysed in order to explain a social or psychological process^5^” (Glaser & Strauss, 1967)

Ethical clearance for the project was obtained both from the “ethical committee the Ministry of Health and Quality Of Life”, Mauritius and the Consultant, “Ministry of Social Security, National Solidarity & Reforms Institutions”. Information about the study was provided to all participants at the Association Alzheimer and subsequently invited to participate. Participation was on voluntary basis and they could leave at any point of time during the study. Information sheet was also provided and relevant details of the project was provided to the patients as well patient’s carer prior to the start.

Consent was obtained through the care giver and the patient. The process and the aim of the study was explained in simple Creole/French language. Verbal consent from both parties (carer and patient) and written consent was obtained from the carer/ care giver and /or the patient

### i. Selection and Description of Participants

All Participants were recruited from the Alzheimer’s Association, at Belle Rose, Mauritius. Sample size was obtained till theoretical saturation reached and no new ideas, data emerged. Interview questions were open ended and clear consisting of 20 open-ended questions. Some instructions were provided for exploring more details, and to metamorphose between topics. The interviewer proceeded to the interview process by engaging the patient in an initial discussion of what are the main types of non-pharmacological therapy (NPT) and how much they were involved. Qualitative research proceeded as mostly-open ended questions with probing.

All interviews were audio recorded and then later transcribed. Person living with Alzheimer’s disease own experiences was written down, noted and recorded with the use of smartphone and later translated from creole or French language to English using google translation. Spoken words and observable behaviour of the individuals formed part in developing a concept. Eventually these were grouped together as particular concepts/themes which generated the field notes.

### ii. Technical Information

Phenomenological interpretative analysis method in qualitative research is an ongoing process of coding data and decoding and grouping similar chunk of data together.

The following steps are required:

Collecting data (Creole language), proceed to coding of data, followed by “categorizing bits of data and then interpreting interview data and writing field notes^6^” (Strauss & Corbin, 1990). “Initial coding involved describing and labelling units of data”. Also “similar kinds of information are grouped together into categories”. Then “relating different ideas and themes to one another^7^” (Rubin and Rubin, 1995). This process was repeated until all raw data are coded once.

This step also involves literature reading (next section; Table 1). Together with the “naïve understanding and the authors’ pre-understanding, these were reflected by using suitable literature to form a comprehensive understanding^8^” (Lindseth & Norberg, 2004). Lastly interpretation takes place.

**Table 1:** The inclusion and exclusion criteria

|  |  |
| --- | --- |
| Inclusion criteria | Population sample: All the participants were recruited from the ‘Alzheimer’s Association’ Centre at Belle Rose, Mauritius. The elderly population aged $\geq 60$ years were included. |
|  | Language: All the participants were local people and creole and French language was commonly used. |
|  | There was no dementia diagnosis. |
|  | No other neuropsychiatric disorders other than AD |
| Exclusion criteria | Population sample: Patients other than those found at the ‘Alzheimer Association’ Centre. Person aged $\leq 59$ years |
|  | Other neuropsychiatry diagnosis other than Alzheimer’s Disease |

### iii. Procedures

No software was used for data processing and analysis in qualitative study. At time of study, there was no exposure to software material for the duration of the research.

A number of studies were searched for online database using Google Scholar and PubMed. Included studies were related to Alzheimer’s disease and dementia of qualitative study methods. The following key terms were searched across all the online databases: “Alzheimer’s Disease”, “Dementia”, “lived-experience”, “dependency”, “emotions”, “quality of life”, “physical exercise”, “group singing”, “music playing”, “family and carer”, “anxiety”, “decision-making”, “interactions with others”, “having a pet”, “outdoor activities”, “awareness into illness”. Table 2 below provides quotations from the individual studies relating them to the main themes and sub-themes in qualitative research.

**Table 2:**
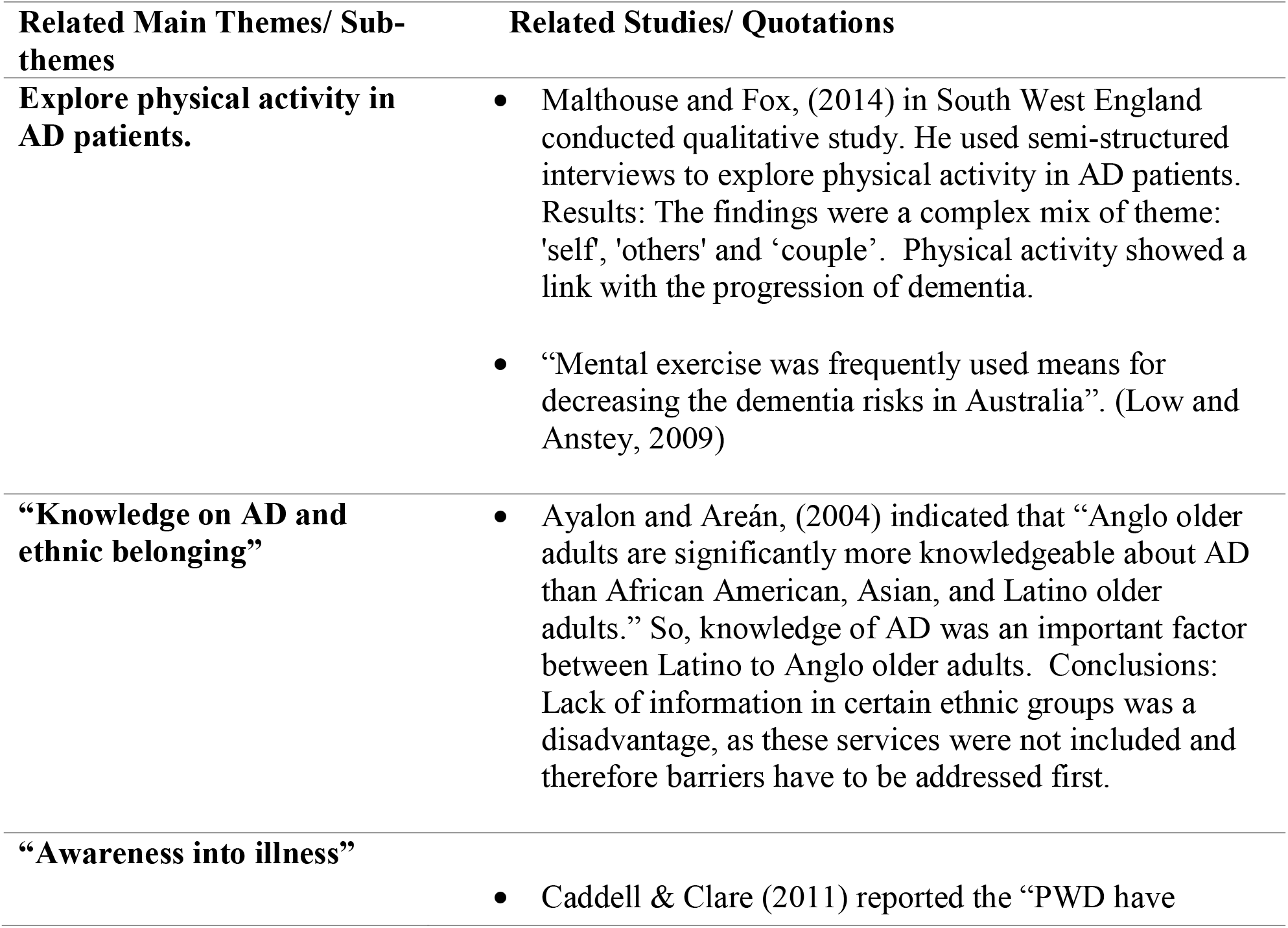
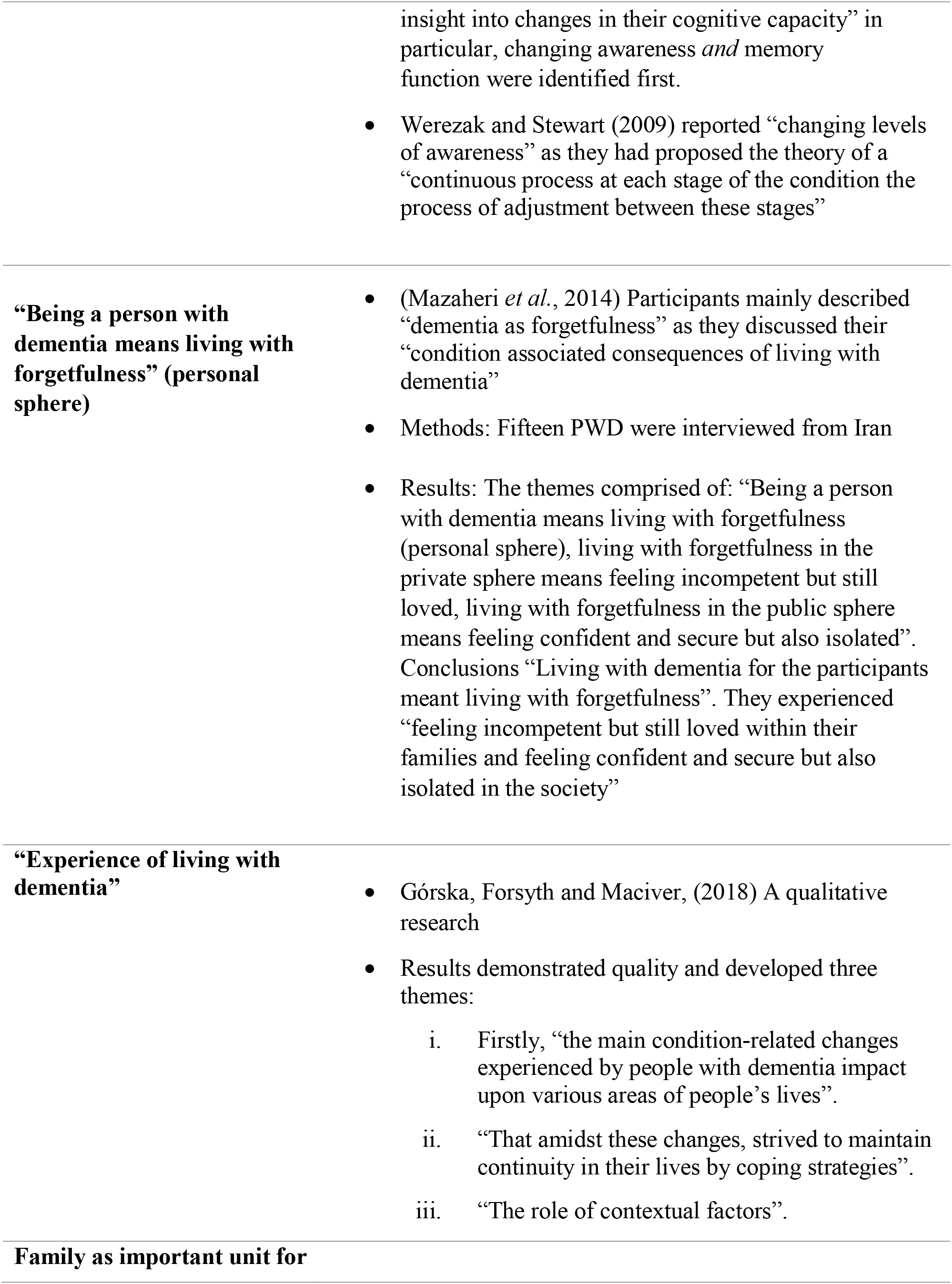
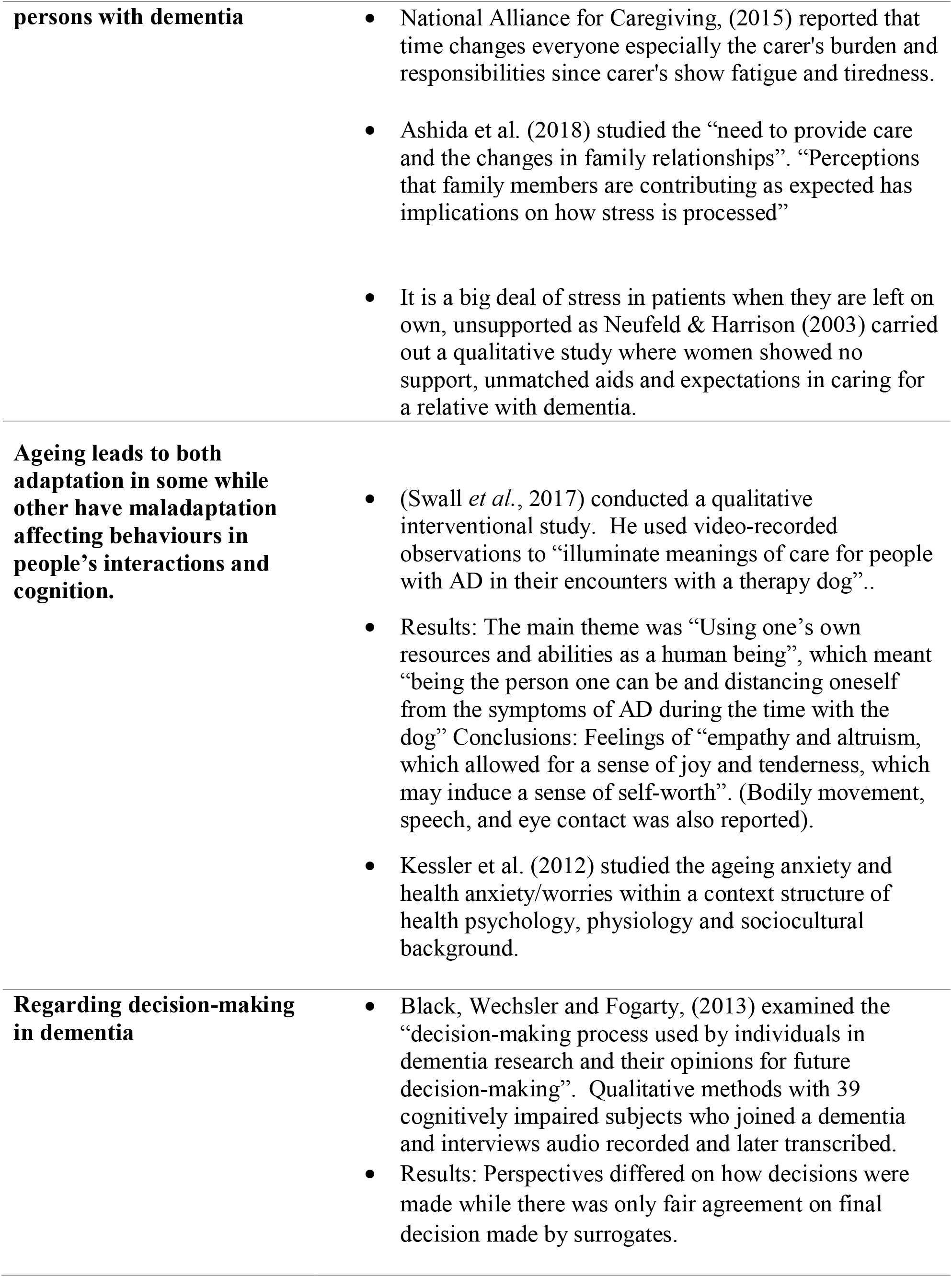
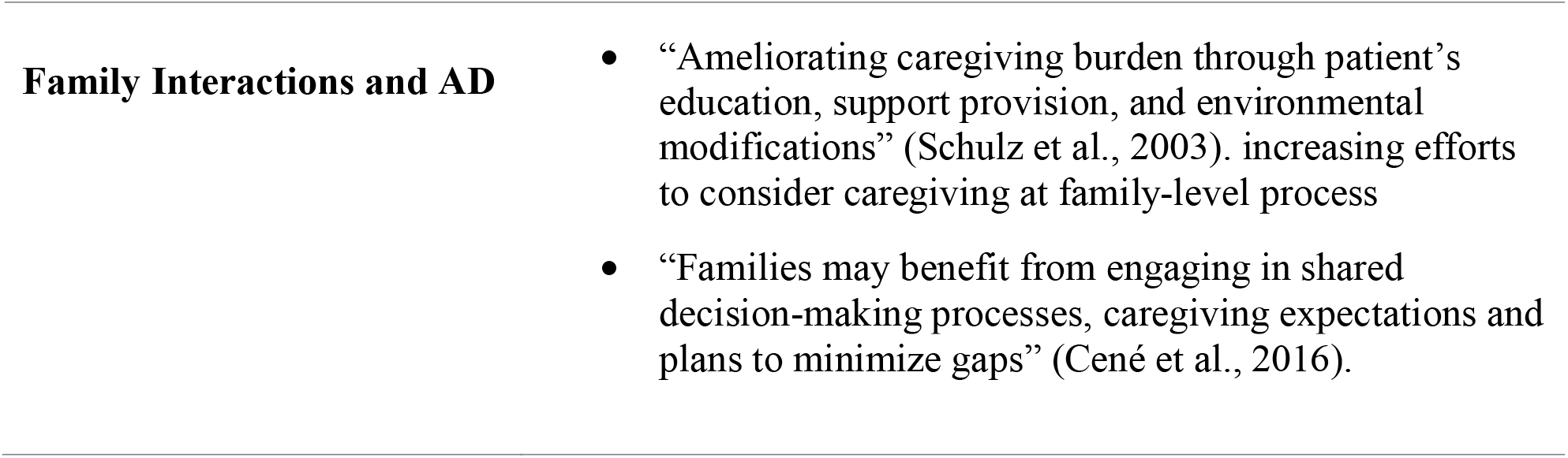
Main themes/ sub-themes and related studies/ quotations

## RESULTS

Results in qualitative study were displayed in the table form and flowcharts. During the process of phenomenological interpretive study, the ‘meaning unit’ has been ‘condensed’ and the main ‘theme’ derived as shown in Table 3, Table 4 below. However, the raw meaning has not been changed into some different meaning. This is known as ‘researcher bracketing’. The “sub-themes”, “themes”, and “main theme” were reflected in literature to form a more proper understanding.

**Table 3:**
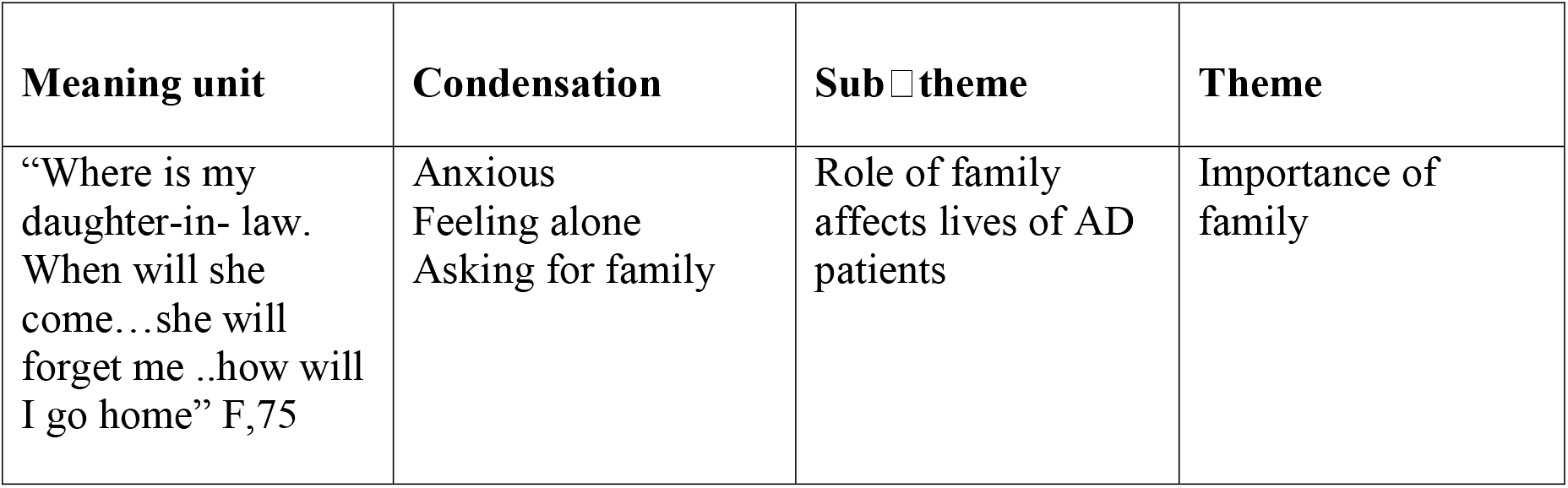
Coding and decoding in qualitative study design

**Table 4:**
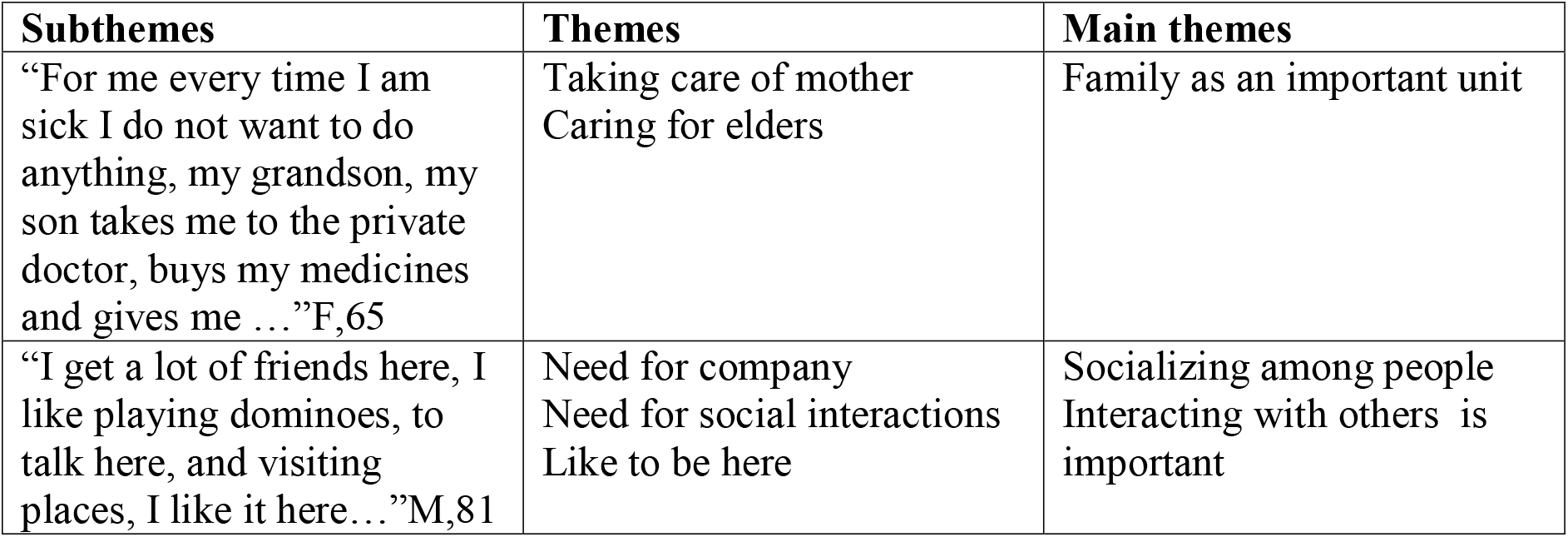
Development of subtheme, themes and main themes in qualitative study

### THEMES

Results of this research emerged as all the themes with similar significance were grouped together and form the following 7 main themes as identified:

- “Active”, “happy”, “friends”
- “I like to sing”, “I can play instrument”
- “I care for the dog”
- “Forget”, “I don’t remember”
- “I am old now”, “I can help”
- “I love my grandchildren” “They take me to the doctor”
- “I can recall”, “old memories”

The flowchart below (Figure 1, Figure 2) depicts the initial set of raw data in condensed form, further analysed and grouped together to demonstrate the role of family as identified by the persons with AD. Here, the emotions were also displayed in association with the important part of qualitative studies.

**Figure 1:**
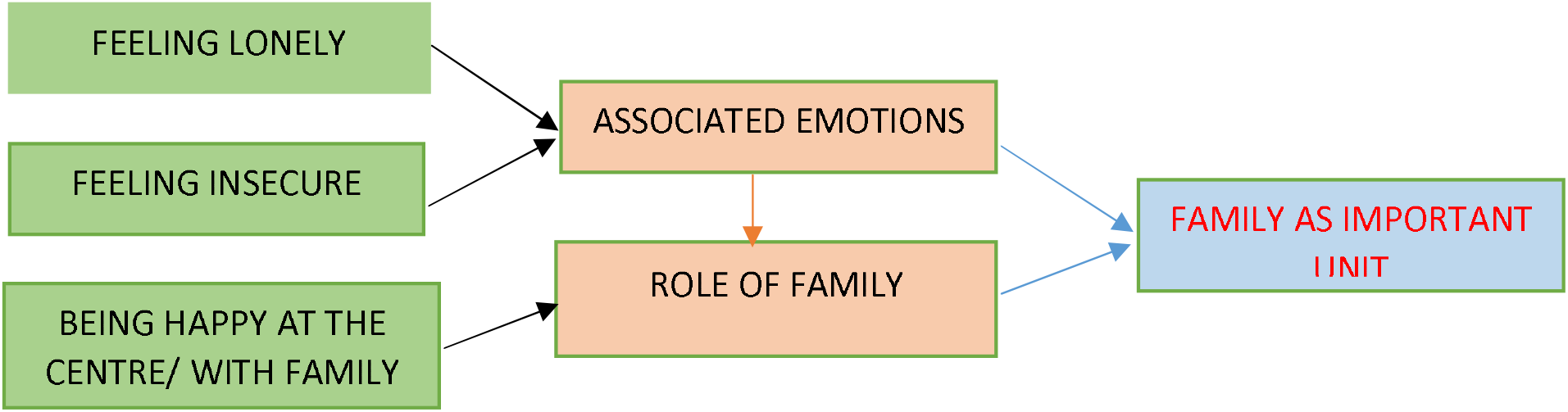
Flowchart showing the coding, the subthemes, and the main themes. The emotions associated with person with AD expressed and grouped together, highlighting the importance of family as NPT.

**Figure 2:**
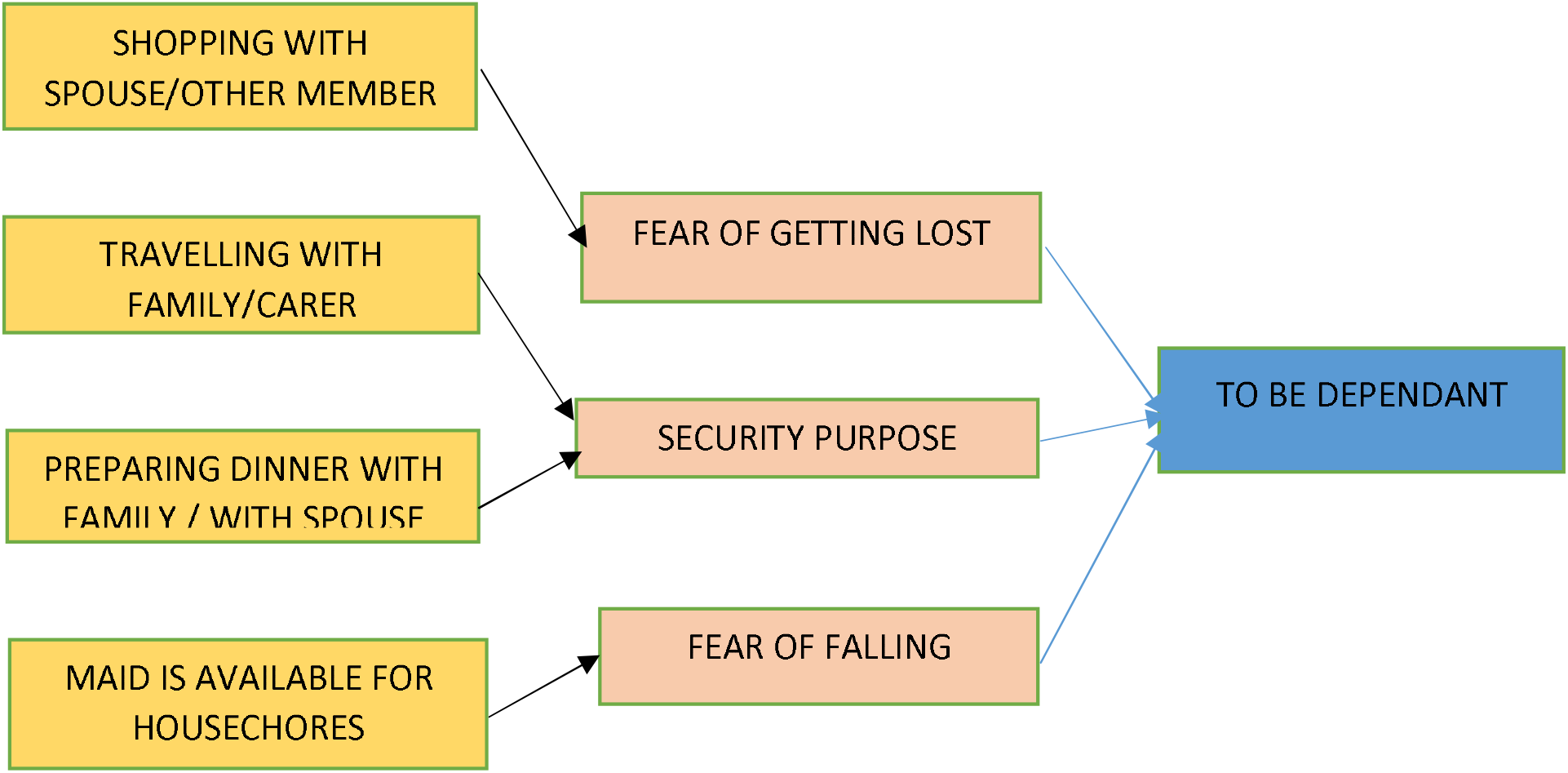
Flowchart showing the coding, the subthemes, and the main themes. The activities of daily living were mostly associated with security of the individual and management of everyday living purposes, which displayed a certain level of being a dependant living person with AD, as qualitative study joins the functional capacity and emotions.

### Theme 1: Participation in activities

Patients talked about their level of activity. Physical activity mentioned by patients were yoga, stretch exercise, breathing exercise. Physical exercise was noted as being important for the patients. It is also a practice of healthy behaviours. Patients have reported that they regularly exercise at the Centre as well as at home.

“I have learnt to do stretch exercise” M, 70. “I do breathing exercise and yoga classes” F, 86.

“I do mild to moderate physical activity like having a walk in the locality area/playground for few minutes…I am active” F, 83.

Others prefer to practice yoga and breathing exercise at home: “one particular days, when I stay at home, in the morning time I do practice yoga and breathing exercises, because we have learnt this from here…”M, 76

### Theme 2: Singing and Music

Music was much appreciated at the Centre especially since it offer music and singing sessions to the elderly. They were active participants also.

“I like singing and I play piano at Church” M, 72. “I like to sing and dance here…the centre has such programmes” F, 64. “I like music, I was a teacher” F, 63. “I can sing with children” F, 68.

### Theme 3: Affection and caring

Caring for pets. Many patients reported that they spent time with their pet at home,

“The dog is like security guard at home… I feed him only once a day, bath the dog” F, 85

“I like birds, they can sing for me” F, 89

### Theme 4: Knowledge on AD

Patient was able to describe their symptoms as loss in personal belonging and forgetfulness symptoms, related to correct educational background on the disease process and manifestation, arising fear in persons with AD.

“‥the disease affects my life as I have loss my key, my wallet” M, 70

“‥I reached the shop and I was standing out there‥,I forgot what to buy…, ‘to forget’ is part of the illness”M,69

### Theme 5: Level of dependency and level of autonomy

Issues relating to ageing and demotivation, low energy levels. Although no physical disability was present, patient was depressed.

“I am old now, I fell tired, I can’t do much” M, 62.

Assistance was required at home for cleaning and cooking as some patients mentioned being more dependent. Some patients have reported that a maid was available to do the cleaning at home and cooking part while others shared the chores.

“I do all the house chores and I am so much tired that my son scolds me all the time‥my house should be clean that’s all I like” F, 69

Furthermore, there was no mention of “being lost” while traveling alone however, they were under care and supervision.

“I usually go to short distances, or take one bus directly to and from home, go to seaside for some time…” M, 65

“‥ I can drive for short distance, to known places, accompanied by my wife,… otherwise I take the bus for long distances and accompanied by a relative…”, M,60

“I am travelling in my daughter’s car accompanied by a driver who drops me to the center, and to supermarket to do shopping and then back home…” F, 79

The persons with AD were never left on their own, while some travelled over short distances either by the bus alone, or a driver would drop them at the center and pick up

### Theme 6: Family support

“I like spending time with family,‥I like spending time with grandchildren, F,82. Describing family as an important unit for support.

One patient reports when I fall sick my children and grandchild takes me to the doctor, they buy my medicine and give me …I love them”F, 86

“I can speak French with my grandchildren, as their mother also does same ‥I am learning too” learning at all phases of life, F,60

One persons with AD showed worriedness and lack of security and repeatedly said: “She will forget me here, how will I go home‥I am afraid, to walk down the lane too many bad guys, what will happen if they hit and snatch my bag‥can you call my daughter-in-law please”, F,60

### Theme 7: Recall memory of the past

AD patients recall their past memories. The long term memory were detailed in long conversations, at times, relating to their childhood and work.

“I listen to mother‥she tells me do this …” F, 87

“I used to do all heavy work, cut cane, take care of my 4 children, cook ‘farata’, I have worked enough…I can travel now and rest, I have married all my sons ‥I am free person” F, 64

AD patients showing derailment with poor recall memory, and say little, not participating or failed to understand the conversation, referring to “the thing” all the times showing aphasia “This centre is for the mentally disturbed patients? I think that’s why I am here, I am mentally sick?” M, 68.

## DISCUSSION

The sampled population was already exposed to both pharmacological and NPT for AD. NPT included the occupational therapy, speech therapy, and behavioural cognitive therapy, reminiscence therapy, group work activities, psychotherapy. These activities engaged the person with AD at the Centre in various ways so the findings in qualitative research was spoken, interactive discussions and interviews that captured the patients with AD lived-experiences. In addition, the main themes “active”, “happy”, “like”, “care” contributed towards the health of the people with AD. As new data emerges it adds to future knowledge. It is important for Mauritius to carry out further research work to focus on NPT and to plan national guidelines and integrated, mixed treatment for AD. Alzheimer’s disease remains an important topic and the patient should be taken care of with multidisciplinary actors and stakeholders acting together to provide evidence-based knowledge and practice.

### Weakness in qualitative research

Interview is a time consuming process. The disease process manifest itself as many symptoms identified as barriers in qualitative research namely:

a. Patients are impatient and don’t like to sit for too long time with strangers
b. They show poor responsiveness towards the researcher
c. They may be aggressive at times
d. Their language problems, agnosia is present
e. Difficulty in understanding instructions (communication problems)
f. Hearing problems, poor vision and age-related problems which limit the accuracy of data

## Data Availability

All data referred to in the manuscript and notes will be made available except patient details and personal information to the publisher

## Footnotes

1 Alzheimer’s Disease International. World Alzheimer Report 2018 The state of the art of dementia research: New frontiers. UK; 2018. 48 p

2 Nelson L, Tabet N et al. *Slowing the progression of Alzheimer’s disease; what works, 2015*;23(Pt B):193-209

3 World Health Organization, top 10 causes of death, 2018 report

4 Glaser B G, Strauss A L 1967 The discovery of Grounded Theory, Aldine, New York

5 Glaser B G, Strauss A L 1967 The discovery of Grounded Theory, Aldine, New York

6 Herbert J. Rubin & Irene S. Rubin, *Qualitative Interviewing (2nd ed.): The Art of Hearing Data,(2^nd^ edn,2005)*

7 Herbert J. Rubin & Irene S. Rubin, *Qualitative Interviewing (2nd ed.): The Art of Hearing Data,(2^nd^ edn,2005)*

8 Lindseth A, Norberg A. et al. *A phenomenological hermeneutical method for researching lived experience, 2004*;18(2):145-53.

